# Language eligibility criteria and enrolled racial and ethnic composition in US clinical trials: a cross-sectional study

**DOI:** 10.64898/2026.10.09.26365106

**Authors:** Alexander M Tolas, Lina Du, Narayan Schutz, Paul Schmiedmayer, Jack W. O’Sullivan, Anders Johnson, Christopher T Longenecker, Ashish Sarraju, Fatima Rodriguez, Eugene Yang, Daniel Seung Kim

**Author notes:** Corresponding Author: Daniel Seung Kim, MD, PhD, MPH 1959 NE Pacific St Box 356422 Seattle, WA 98195.

## Abstract

**BACKGROUND:** Federal policy now requires sponsors to submit diversity action plans for clinical trials, yet the modifiable features of trial design associated with enrolled composition remain poorly characterized. English-language proficiency requirements are written into eligibility criteria and recorded as free text in a public registry, making them visible and, in principle, amenable to revision, but their relationship to enrolled composition has been examined only within single disease areas and in small samples of trials. We examined whether these requirements and documented language accommodation were associated with the racial and ethnic composition of enrolled participants.

**METHODS AND FINDINGS:** We conducted a cross-sectional study of interventional trials registered on <u>ClinicalTrials.gov</u> with at least one US site and posted results, using a registry snapshot retrieved in October 2026. The primary cohort comprised the 44,487 trials with exclusively US sites, of which 21,924 (49.3%) reported baseline race and 17,434 (39.2%) reported baseline ethnicity with counts that reconciled with the posted denominator. Both exposures were classified from free-text eligibility criteria using regular expressions validated against independent physician review of 150 trials. For the English-language requirement classifier, sensitivity was 88.2% and specificity was 99.2%. The outcome was the trial-level proportion of enrolled participants in each category, analyzed using generalized linear models adjusted for intervention class, therapeutic area, sponsor, phase, enrollment, sites, start year, US Census region, and the population composition of the trial’s site states. An English-language proficiency requirement appeared in 5,736 trials (12.9%); its prevalence rose from 7.4% of trials starting in 2010 to 26.2% in 2023 (odds ratio [OR] per decade 2.91, P<0.001), whereas documented accommodation, an eligibility statement allowing participation in a language other than English (Spanish in 99.9% of flagged trials), remained rare (788 trials, 1.8%). The requirement was associated with lower enrolled proportions of Asian (OR=0.85, 95% CI 0.77-0.93, *P*=0.001) and Hispanic/Latinx participants (OR=0.85, 95% CI 0.79-0.91, *P*<0.001), with no evidence of an association for White participants (OR=0.99, 95% CI 0.95-1.04, *P*=0.75). Documented accommodation was associated with a higher proportion of Hispanic/Latinx participants (OR=3.23, 95% CI 2.82-3.71, *P*<0.001). Geographic adjustment did not change the statistical significance of the requirement associations and attenuated the accommodation association. The main limitations were the cross-sectional design, which precludes causal inference; the restriction of enrollment analyses to trials reporting baseline demographics, which were more recent than those without such reporting; and possible misclassification of automatically classified exposures.

**CONCLUSIONS:** English-language proficiency requirements became more than three times as common between 2010 and 2023 and were associated with lower enrolled proportions of Asian and Hispanic/Latinx participants. Less common documented language accommodation was associated with a higher enrolled proportion of Hispanic/Latinx participants. Because these criteria are investigator-written and publicly recorded, they may be a tractable target for improving enrollment equity, although whether revising them changes who enrolls requires prospective evaluation.

## INTRODUCTION

The number of US residents who speak a language other than English at home nearly tripled between 1980 and 2019, reaching 67.8 million (nearly 20% of the US population)[1]. Roughly 8% of US residents aged 5 years or older speak English less than “very well”[1,2]. A non-English-language preference is associated with poorer cancer care[3], worse cancer-related outcomes[4], and poorer recovery after cardiac surgery[5]. Professional interpreters narrow these gaps[6,7], and their absence is associated with increased clinical errors[8].

Language also influences participation in clinical trials. Patients who do not speak English are offered trial enrollment less often, consent less often, and complete trials less often[9–11]. Among the 21 research centers receiving the most National Institutes of Health funding in 2018, institutional review board policies on consenting non-English speakers were inconsistent, leaving standards subject to local practice variation[12]. Fewer than 3% of trials offer translation, and roughly 19% require English-language proficiency[13]. The evidence supporting drug and device approval therefore comes from populations that may differ from those who receive the therapies after approval.

The Food and Drug Omnibus Reform Act of 2022 requires sponsors of pivotal trials to file diversity action plans naming subgroup enrollment goals and the means of reaching them[14,15]. Language-related eligibility criteria are a potential target for improving enrollment equity. Egleston *et al* first characterized trials requiring English-language proficiency in 2015, reporting a rise from 1.7% before 2000 to 9.0% after 2010, driven by behavioral trials[16]. Muthukumar *et al* subsequently found that, of 14,367 registered trials, 18.98% (N=2,727) required English-language proficiency, while 2.71% (N=390) documented any non-English accommodation[17]. The same pattern has since been reported in pediatric[18], gynecologic oncology[19], and cardiovascular research[20], and in studies of language-access practices at academic medical centers[21], while other studies have described the logistics and costs of translation and interpretation in trials[22,23]. Two studies have compared enrolled composition according to language eligibility criteria within a single disease area[24,25], but none has tested the association registry-wide, across therapeutic areas, with covariate adjustment and control exposures.

Sponsors and reviewers have promoted digital and decentralized designs as instruments of inclusion, because remote participation removes travel and scheduling barriers[26–28]. However, any application or text-message protocol must be built and validated in a particular language, raising costs[29]. In cardiovascular and cardiometabolic digital trials, we previously found that more than half required English-language proficiency for enrollment[20]. The present study extends that work registry-wide and examines the association between the English-language proficiency requirement and enrolled racial and ethnic composition.

We therefore used structured baseline race and ethnicity tables submitted through mandatory results reporting on <u>ClinicalTrials.gov</u> to determine whether English-language proficiency requirements are associated with lower enrolled proportions of racial and ethnic minority groups in US-based clinical trials. Separately, we sought to determine whether documented language accommodation was associated with higher proportions of racial and ethnic minority groups. Finally, we investigated whether digital or decentralized trials were more likely to impose English-language requirements than conventional trials, and whether these modalities were associated with differences in enrolled racial and ethnic composition.

## METHODS

### Ethics

This study used publicly available, aggregate, and de-identified registry data. It was reviewed by the University of Washington Institutional Review Board and determined not to constitute human subjects research.

### Design and data source

We analyzed trial-level records retrieved in October 2026 from the <u>ClinicalTrials.gov</u> application programming interface (API, version 2), requesting interventional studies with at least one US site and posted results. The registry has accepted registrations since February 2000; trials in the analytic cohort had start dates from 2000 through 2025, and 4 trials (0.01%) had no recorded start year and were excluded from year-stratified analyses. Cohort construction proceeded in three stages (see **Supplemental Figure 1**), following the STROBE guidelines for cross-sectional studies[30]. A total of 56,544 interventional trials with at least one US site and posted results were identified. Of these, we retained only trials with exclusively US sites (N=44,487), because trials enrolling participants abroad draw from populations with different demographic compositions. Finally, we required baseline race or ethnicity data, which yielded 21,924 trials for race and 17,434 trials for ethnicity.

### Exposures

We classified English-language proficiency requirements and documented language accommodation from the free-text eligibility criteria field using case-insensitive regular expressions (see **Supplemental Appendix** for verbatim classifier patterns and validation). The classifier for the English-language proficiency requirement matched constructions denoting ability, fluency, proficiency, literacy, or native use of the English-language, together with their negated forms. Because the field concatenates inclusion and exclusion criteria text, the classifier captures either English-language proficiency as an inclusion criterion or lack of proficiency as an exclusion criterion, without distinguishing between them. Constructions that negate the requirement itself, such as “English fluency is not required”, would be misclassified as imposing a requirement; we searched for them at the level of the matching sentence and found none among exposed trials, so this source of misclassification did not arise in this corpus.

The documented language-accommodation classifier matched explicit two-language allowances, such as “English or Spanish” and “English or [insert language]-speaking”. We use “documented language accommodation” rather than “bilingual accommodation” because the classifier identifies only accommodation recorded in the registry’s eligibility text, which may understate what trials provide in practice. It also misses accommodation recorded elsewhere in the <u>ClinicalTrials.gov</u> record and trials listing three or more languages in comma-separated form.

Two further exposures were defined as controls. First, we identified any mention of “Spanish” in the eligibility criteria field, which we expected to reproduce the association seen with Hispanic/Latinx enrollment (positive control). The accommodation classifier matches a bilingual eligibility statement and therefore does not capture interpreter availability or translated study materials, which are recorded outside the eligibility criteria field; fewer than 2% of flagged trials mention an interpreter and fewer than 1% mention translation. Second, we identified the eligibility requirement of “must be able to take oral medications”, which we expected to have no association with enrolled racial or ethnic composition because it specifies a functional requirement unrelated to race, ethnicity, or language (negative control). Because every trial classified as documenting accommodation also mentions Spanish, the accommodation exposure is nested within the Spanish-mention exposure, so the two are not independent; we therefore also estimated the association in the disjoint subset of trials that mention Spanish without being classified as accommodating, excluding every accommodation-classified trial from that analysis so the comparator comprised trials that neither mention Spanish nor meet the accommodation definition.

We identified digital or decentralized trials by searching the title and summary text for device-, monitoring-, application- and telehealth-oriented terms (see **Supplemental Appendix** for full terms searched).

To validate the classifier, we drew a random sample of 150 trials from the 56,544 harvested records. Two board-certified physicians in Internal Medicine independently adjudicated each trial’s eligibility criteria, blinded to the classifier label, and answered whether participation was conditioned on the participant’s ability to use English proficiently. The physicians agreed on 147 of 150 trial’s eligibility classifications (98.0%; Cohen’s *κ*=0.908, 95% CI, 0.80-1.0). This yielded a sensitivity of 88.2% (15/17), specificity of 99.2% (132/133), positive predictive value of 93.8% and negative predictive value of 98.5% (see **Supplemental Table 1**). At the 5% prevalence of some classified exposures, a random sample of this size yields few exposed records, so the precision of sensitivity is limited; for the modality classifier we therefore drew a sample stratified on the classifier label, which at 150 classifier-positive records gives a standard error on sensitivity of approximately 0.02 rather than 0.10.

We additionally validated the documented language accommodation classifier against the same 150-trial random sample. The two physician reviewers had 100% agreement (Cohen’s *κ*=1.00), agreeing on all 150 reviewed trials, and the automated regular expression classifier had a sensitivity of 85.7% (6/7), specificity of 100% (143/143), a positive predictive value of 100%, and a negative predictive value of 99.3%. The digital or decentralized modality classifier was validated separately on a sample of 300 trials stratified on the classifier label, 150 positive and 150 negative, adjudicated with reviewers blinded to the classifier label using the brief title, brief summary, and intervention names. The term list combining application, telehealth and decentralized constructions with named wearable devices had a positive predictive value of 86.5% (95% CI, 78.2%-91.9%) and a specificity of 99.4%. A broader list that included the unqualified term “monitoring” had a positive predictive value of only 66.7%, and the modality exposure was therefore restricted to the narrower list.

### Outcomes, covariates, and analysis

All analyses were performed using Python 3.11 with pandas, statsmodels, and scipy.

Outcomes of interest were trial-level proportions of enrolled participants in each racial and ethnic category, taken from the baseline characteristics module of the <u>ClinicalTrials.gov</u> API, which reports race and ethnicity as separate measures with fixed vocabularies. The primary outcomes were the proportion of Asian participants and, separately, the proportion of Hispanic/Latinx participants. Proportions in other categories (e.g., White and Black participants) were analyzed as secondary outcomes hypothesized a priori to be null. The secondary estimates were interpreted in light of the compositional structure of the race data; ethnicity was analyzed separately.

Because the outcome is a trial-level proportion of participants in a racial or ethnic category (rather than a per-participant binary indicator), we used binomial generalized linear models with a logit link[31], which yielded odds ratios (ORs) describing associations with the odds corresponding to a trial’s enrolled proportion in a category of interest. We used heteroskedasticity-robust (White/HC0) standard errors throughout[32]. A trial entered a given model when the registry posted a count for that category, so the three race models differ slightly in size: 20, 19 and 25 trials of the 21,924 in the race cohort lacked an Asian, White or Black count, respectively, and 18 of the 17,434 in the ethnicity cohort lacked a Hispanic/Latinx count. No trial was excluded from a race or ethnicity model for missing covariate data. Five trials were excluded from the modality model for missing enrollment or start year. Adjusted models included intervention class (behavioral, drug/biologic/device, or other), therapeutic area (eight categories, listed in the **Supplemental Appendix**), industry sponsorship, presence of a registered clinical phase, total enrollment, total number of sites, and trial start year. Adjusted models additionally included US Census region of the trial’s sites and the population share of the outcome-matched group across the states containing trial sites, computed from the 2023 American Community Survey five-year estimates (table B03002) and weighted by state population, to account for the possibility that trials imposing language criteria are located in less diverse areas. Site states were deduplicated before this calculation, so a state hosting several sites counted once; 28.4% of trials listed sites in more than one state, and trials whose states fell in more than one census region were assigned a distinct Multiregion category rather than excluded. No trial in the primary cohort lacked a parseable site state.

We modeled the trend in each exposure’s prevalence over time using a binomial generalized linear model with a logit link[31], regressing the trial-level binary exposure indicator (English-language proficiency requirement or documented language accommodation) on trial start year, treated as a continuous predictor. The model was unadjusted for other covariates and used HC0 standard errors, as with the primary models[32]. It was fitted over the full harvested cohort with a recorded start year (N=56,540).

Multiple testing was addressed using Benjamini-Hochberg false discovery rate (FDR) corrections[33] over a declared family of 73 tests, comprising the 64 comparisons tabulated in **Supplemental Table 2** together with the four trend functional-form comparisons, the disjoint Spanish-only analysis, and the four exposure-by-sponsor interactions. All P values reported in the text are FDR-adjusted over that family. Adjusting over the 64 tabulated comparisons alone changes no reported conclusion.

Each OR describes the composition of a trial’s enrolled participants rather than any individual’s chance of enrolling; in the Asian-enrollment model, an OR of 0.85 indicates 15% lower ods corresponding to the enrolled Asian proportion in trials requiring English-language proficiency, conditional on the included covariates. Models were unweighted, so the estimates describe the average trial rather than the average participant.

Two hypotheses were primary: that English-language proficiency requirements for enrollment would be associated with lower enrolled proportions of Asian and Hispanic/Latinx participants, and that documented language accommodation would be associated with a higher enrolled proportion of Hispanic/Latinx participants. Controls were specified as described above: a positive control exposure (any mention of “Spanish”), a null exposure (ability to take oral medications), and a null outcome (the enrolled proportion of White participants). Intervention type was examined as an exploratory sensitivity analysis.

## RESULTS

### Cohort and trial characteristics

Of 56,544 US interventional trials with posted results, 44,487 had exclusively US sites and formed the primary analytic cohort. Of these trials, 21,924 had race data and 17,434 had ethnicity data (see **Supplemental Figure 1**). English-language proficiency was required by 5,736 trials (12.9%). Documented language accommodation was less common, recorded in 788 trials (1.8%).

Trials requiring English-language proficiency differed on several characteristics (summarized in **Table 1**): they were less often industry-sponsored (8.7% vs 27.9%), less likely to have a registered phase (30.8% vs 66.4%), and more recent (median start year, 2018 vs 2014). They were also more often behavioral (37.4% vs 7.8%) or digital (11.9% vs 2.3%).

**TABLE 1.** Characteristics of US interventional trials with posted results, by English-language proficiency eligibility requirements (N=44,487).

| **Characteristic** | **English-language proficiency required (N=5,736)** | **No documented English-language proficiency requirement (N=38,751)** | **Overall (N=44,487)** |
| --- | --- | --- | --- |
| Industry sponsor, % | 8.7 | 27.9 | 25.5 |
| Has clinical phase, % | 30.8 | 66.4 | 61.8 |
| Behavioral intervention, % | 37.4 | 7.8 | 11.6 |
| Drug/biologic/device, % | 50.1 | 85.0 | 80.5 |
| Digital/decentralized, % | 11.9 | 2.3 | 3.6 |
| Median start year | 2018 | 2014 | 2015 |
| Oncology, % | 10.8 | 24.5 | 22.7 |
| Mental/behavioral health, % | 18.2 | 7.2 | 8.6 |
| Cardiometabolic, % | 11.5 | 10.5 | 10.7 |
| Infectious, % | 5.0 | 6.0 | 5.9 |

### Temporal trends in English-language proficiency requirements

English-language proficiency requirements increased beyond the period covered by the prior registry-wide estimate[16], from 7.4% of trials starting in 2010 to 26.2% in 2023 (OR per year=1.11, 95% CI, 1.11-1.12, P<0.001; OR per decade=2.91; N=43,614 trials with a start year from 2000 through 2023; see **Figure 1** and **Supplemental Table 3**). Trials with start years after 2023 are reported in **Supplemental Table 3** but excluded from the trend estimate, as results-reporting lag leaves them incompletely captured. Documented language accommodation also rose in relative terms (OR per decade=2.59, P<0.001), but stayed below 4% every year. The prevalence of digital or decentralized trials rose in parallel, from 1.1% to 11.6%. Neither a quadratic term nor a four-degree-of-freedom spline improved on the log-linear fit (P=0.34 and P=0.29), so these comparisons provided no evidence of acceleration, although year-specific effects did fit the data better than the linear term (P=0.008), indicating variation not captured by the linear trend.

**FIGURE 1.**
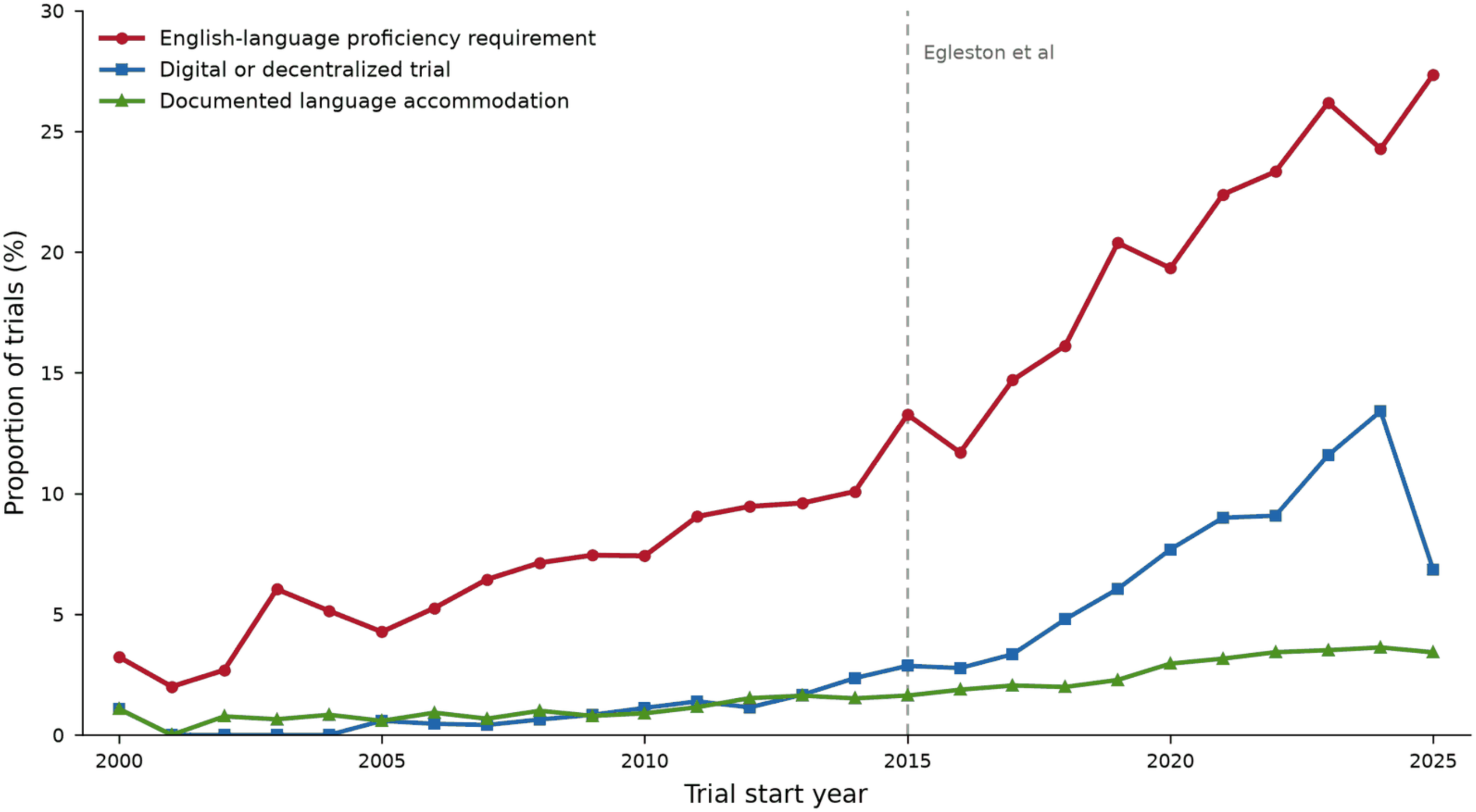
Prevalence of English-language proficiency eligibility requirements, documented language accommodation, and digital/decentralized modality among US interventional trials with posted results, by trial start year (2000-2025).

### Language eligibility criteria and enrolled composition

Trials requiring English-language proficiency enrolled a significantly lower proportion of Asian participants (OR=0.85, 95% CI, 0.77-0.93, P=0.001, N=21,904, see **Table 2** and **Figure 2**). The requirement was also associated with a lower enrolled proportion of Hispanic/Latinx participants (OR=0.85, 95% CI, 0.79-0.91, P<0.001, N=17,416). There was no evidence of an association with the enrolled proportion of White participants (OR=0.99, 95% CI, 0.95-1.04, P=0.75, see **Table 2**). The enrolled proportion of Black participants was slightly higher (OR=1.08, 95% CI, 1.02-1.15, P=0.02 after Benjamini-Hochberg correction). Because race proportions within a trial sum too one, the race estimates are interdependent; Hispanic/Latinx ethnicity is reported separately. In an additional compositional analysis, using White participants as the reference category, there was no evidence of difference in the Black-to-White ratio (ratio of ratios 1.02, 95% CI, 0.96-1.08, P=0.60), whereas the Asian-to-White ratio was lower (0.93, 95% CI, 0.89-0.98, P=0.009).

**FIGURE 2.**
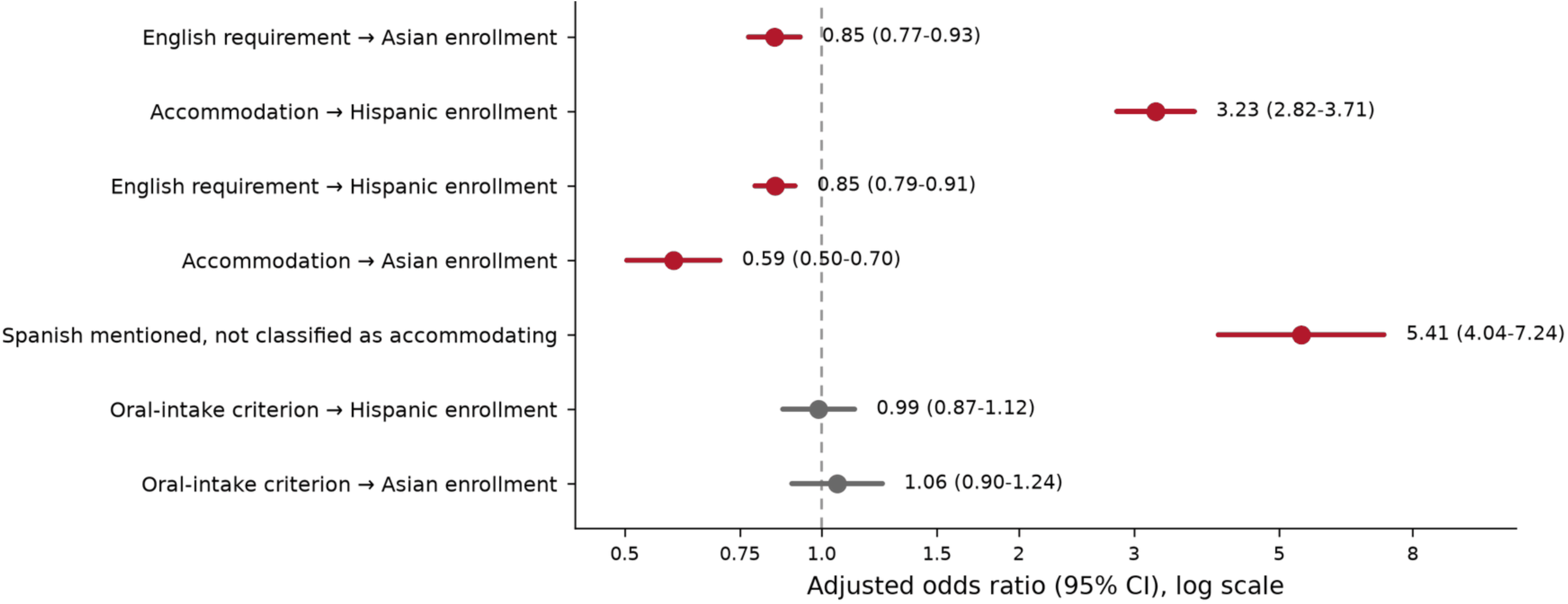
Adjusted odds ratios (OR) for associations of English-language proficiency requirements and documented language accommodation with enrolled racial and ethnic minority group composition. Points are ORs, and horizontal lines are 95% confidence intervals; the vertical reference line marks the null. Rows appear in four groups, in order: the two pre-specified primary hypotheses (English-language proficiency requirement and Asian enrollment; documented language accommodation and Hispanic/Latinx enrollment); two secondary associations (the requirement and Hispanic/Latinx enrollment; accommodation and Asian enrollment); the positive control exposure (a Spanish mention in the eligibility criteria field among trials not classified as accommodating); and the negative control exposure (an oral-intake eligibility criterion), for each primary outcome. Red marks indicate FDR-adjusted P<0.05.

**TABLE 2.** Primary and companion adjusted models. Odds ratios (OR) from binomial generalized linear models with logit link and robust standard errors, adjusted for intervention class, therapeutic area, industry sponsorship, phase, log enrollment, log sites, mean-centered start year, US Census region, and the population share of the outcome-matched group across states containing trial sites.

| Model | Outcome | Exposure | OR (95% CI) | N Trials | N Trials with Exposure | FDR-adjusted *P* |
| --- | --- | --- | --- | --- | --- | --- |
| %Asian-English | %Asian in enrolled trials | English Required | 0.85 (0.77-0.93) | 21,904 | 3,397 | 0.001 |
| %Hispanic-English | %Hispanic/Latinx in enrolled trials | English Required | 0.85 (0.79-0.91) | 17,416 | 2,765 | <0.001 |
| %White-English | %White in enrolled trials | English Required | 0.99 (0.95-1.04) | 21,905 | 3,398 | 0.75 |
| %Black-English | %Black in enrolled trials | English Required | 1.08 (1.02-1.15) | 21,899 | 3,397 | 0.02 |
| %Asian-Oral Rx | %Asian in enrolled trials | Able to swallow medications (negative control) | 1.06 (0.90-1.24) | 21,904 | 718 | 0.55 |
| %Hispanic-Oral Rx | %Hispanic/Latinx in enrolled trials | Able to swallow medications (negative control) | 0.99 (0.87-1.12) | 17,416 | 590 | 0.86 |
| %Hispanic-accommodation | %Hispanic/Latinx in enrolled trials | Documented language accommodation | 3.23 (2.82-3.71) | 17,416 | 412 | <0.001 |
| %Asian-accommodation | %Asian in enrolled trials | Documented language accommodation | 0.59 (0.50-0.70) | 21,904 | 393 | <0.001 |
| %White-accommodation | %White in enrolled trials | Documented language accommodation | 0.64 (0.57-0.71) | 21,905 | 394 | <0.001 |
| %Black-accommodation | %Black in enrolled trials | Documented language accommodation | 1.11 (0.98-1.26) | 21,899 | 393 | 0.14 |
| %Hispanic-"Spanish" | %Hispanic/Latinx in enrolled trials | Mention of "Spanish" in the eligibility criteria field | 3.78 (3.33-4.30) | 17,416 | 533 | <0.001 |

Trials documenting language accommodation enrolled a significantly higher proportion of Hispanic/Latinx participants (OR=3.23, 95% CI, 2.82-3.71, P<0.001, N=17,416). Documented language accommodation was also associated with lower enrolled proportions of Asian (OR=0.59, 95% CI, 0.50-0.70, P<0.001, N=21,904) and White participants (OR=0.64, 95% CI, 0.57-0.71, P<0.001), but there was no evidence of a difference in the enrolled proportion of Black participants (OR=1.11, 95% CI, 0.98-1.26, P=0.14). Because race categories within a trial are compositional, a higher enrolled share of one category is necessarily offset by lower shares of others, so the race estimates are interdependent. Ethnicity is reported separately, with its own denominator; the Hispanic/Latinx estimate is therefore not constrained by the distribution of race categories, although ethnicity proportions are themselves compositional.

Compared with the 2020 US Census population shares, participant-weighted enrollment shares were 0.58 times the population share for Asian participants and 0.78 times for Hispanic/Latinx participants. In contrast, Black (1.29) and White (1.15) participants had participant-weighted enrollment shares at or above their population shares (see **Supplemental Figure 2**). Geographic adjustment left associations with English-language proficiency requirements materially unchanged, but attenuated those with documented language accommodation: across four specifications ranging from no geographic adjustment to adjustment for both US Census region and site-state population share, the English-language requirement association with Asian enrollment changed from OR=0.86 (95% CI, 0.78-0.94) to OR=0.85 (95% CI, 0.77-0.93), and the accommodation association with Hispanic/Latinx enrollment from OR=4.11 (95% CI, 3.59-4.70) to OR=3.23 (95% CI, 2.82-3.71) (see **Supplemental Table 4**).

### Control analyses

The alternative exposures identified as positive and negative controls behaved as predicted. Mentions of “Spanish” in the eligibility criteria field were also associated with a higher enrolled proportion of Hispanic/Latinx participants (OR=3.78, 95% CI, 3.33-4.30, P<0.001). The accommodation exposure is nested within the positive control Spanish-mention exposure, since every trial classified as accommodating also mentions Spanish; in the disjoint subset of 121 trials that mention Spanish without being classified as accommodating, compared with the 16,900 trials meeting neither definition and excluding all 413 accommodation-classified trials, the point estimate was larger (OR=5.41, 95% CI, 4.04-7.24, P<0.001). The eligibility requirement of being able to take oral medications showed no evidence of an association with Asian (OR=1.06, 95% CI, 0.90-1.24, P=0.55) or Hispanic/Latinx enrollment (OR=0.99, 95% CI, 0.87-1.12, P=0.86; see **Table 2** and **Figure 2**).

### Exploratory analyses by trial characteristics

We explored whether the association was stronger in trials in which English-language comprehension may be functionally required. In exploratory sensitivity analyses, the inverse association between English-language proficiency requirements and the enrolled proportion of Asian participants was larger in magnitude in behavioral trials than in drug/biologic/device trials, where it was not significant; the interaction was significant after adjustment for multiplicity (OR=0.77, 95% CI, 0.62-0.94, P=0.03, see **Supplemental Figure 3**). The enrolled proportion of Hispanic/Latinx participants was significantly lower in both behavioral and drug/biologic/device trials requiring English-language proficiency, with no evidence of interaction (OR=0.91, 95% CI, 0.78-1.06, P=0.26).

**Supplemental Figure 4** presents associations stratified by therapeutic area and sponsor type. The direction of association was generally consistent: the English-language proficiency requirement was associated with lower Asian enrollment in all eight therapeutic areas (ORs, 0.65-0.94) and with lower Hispanic/Latinx enrollment in seven of eight (ORs, 0.61-1.44), and documented language accommodation was associated with higher Hispanic/Latinx enrollment in every area (ORs, 1.74-5.14) and lower Asian enrollment in all eight (ORs, 0.49-0.87). Within non-industry trials, all four exposure-outcome associations were significant after adjustment for multiplicity, whereas none of the four was significant within industry-sponsored trials. The ratio of the separately fitted stratum-specific odds ratios for documented language accommodation and Hispanic/Latinx enrollment was 2.1 (non-industry OR=3.35, 95% CI, 2.91-3.85; industry OR=1.59, 95% CI, 0.79-3.20); the industry estimate is imprecise and did not establish an absence of association. The formal exposure-by-sponsor interaction, which constrains the coefficients of other covariates to be common across sponsor groups, was significant for documented language accommodation with both Hispanic/Latinx enrollment (ratio of ORs 0.36, 95% CI, 0.20-0.66, FDR P=0.002) and Asian enrollment (2.18, 95% CI, 1.23-3.87, FDR P=0.02), but was not significant for the English-language requirement with either outcome.

Digital or decentralized trials had higher adjusted odds of imposing English-language proficiency requirements (OR=1.93, 95% CI, 1.70-2.19, P<0.001, N=44,482). There was no evidence, however, that these modalities were associated with differences in enrolled proportions of White, Black, Asian, or Hispanic/Latinx participants (see **Supplemental Figure 5**).

## DISCUSSION

In 44,487 US interventional trials with posted results, English-language proficiency requirements were associated with lower enrolled proportions of Asian and Hispanic/Latinx participants, while documented language accommodation was associated with a higher enrolled proportion of Hispanic/Latinx participants. Three findings are consistent with a language-based mechanism: first, there was no evidence of association between English-language requirements and the enrolled proportion of White participants; the inverse association with Asian enrollment was stronger in behavioral trials, where English-language comprehension is functionally necessary; and Spanish mentions were associated with Hispanic/Latinx enrollment, whereas oral-medication controls showed no evidence of an association with either primary outcome.

### Relation to prior work

This study extends prior prevalence studies by examining enrolled racial and ethnic composition. More than a decade of research has counted language requirements with increasing precision: Egleston *et al* described the trend and its correlates[16], Muthukumar *et al* established registry-wide prevalences of 18.98% of English-language proficiency requirements and 2.71% documented language accommodation[17], and domain-specific studies have found the same pattern in pediatric[18], gynecologic oncology[19], and cardiovascular trials[20]. Smaller studies have connected a language criterion to enrolled composition within a single disease area[24,25], but none has examined this association across the registry and across therapeutic areas in relation to the outcome these criteria are presumed to affect (lower enrollment of racial and ethnic minorities in trials[34,35]), which mandatory results reporting now makes feasible[36].

Our findings also complement the literature on barriers to trial participation. Unger *et al* showed that structural and clinical barriers, including eligibility, outweigh patient reluctance[9], and that patients consent at high rates when they are offered trial enrollment[10]. Language criteria act before consent, at the point of eligibility, and thus constitute a structural barrier[37,38].

Language criteria differ from comorbidity-based exclusions in ways relevant to policy. Excluding patients with organ dysfunction often has a defensible safety rationale. However, excluding those who do not speak English fluently rarely does, outside of trials with language-dependent endpoints. Efforts to broaden eligibility criteria have so far concentrated on comorbidity and organ function[39,40]. Our results, which link English-language proficiency requirements to lower enrolled proportions of Asian and Hispanic/Latinx participants, suggest that language requirements also warrant consideration in efforts to broaden eligibility criteria.

### The digital inclusion paradox

Digital health and decentralized trials are often intended to widen access because remote participation removes travel and scheduling barriers[26–28]. We found that digital trials had higher adjusted odds of imposing an English-language proficiency requirement (OR=1.93), consistent with our prior finding that over half of cardiovascular digital trials required English-language proficiency for participation[20]. Digital designs may therefore retain language barriers, even as they reduce travel barriers, although language models may lower the cost of producing translations for review by trained medical translators[29]. Reviews of decentralized trials already list digital literacy and device access among the risks to equity[41,42]. Language access warrants similar attention, and reports of large siteless trials should state which languages the platform supported.

### Policy implications

Sponsors of pivotal trials must now file diversity action plans naming enrollment targets and the means for reaching them[14,15]. Most correlates of poor trial diversity are structural and slow to change. Language criteria differ in three respects: they are explicit, appearing as text in a public registry field; they act directly on who may enroll rather than through less direct pathways, such as referral or trust in the medical establishment; and, being written by investigators and publicly recorded, they are in principle amenable to revision, although the scientific justification for any individual criterion was not assessed here.

Documenting language accommodation may offer another actionable target. Documented language accommodation appeared in a small minority of trials and was strongly associated with a higher enrolled proportion of Hispanic/Latinx participants. The magnitude of this association and its concordance with the Spanish-mention exposure support further evaluation of language accommodation as a means of improving recruitment, although targeted recruitment may contribute to both associations. Requiring sponsors to state whether patients with a non-English-language preference are eligible, and, if not, why, would make the criterion visible with limited additional reporting, as others have urged[13].

### Limitations

This is a cross-sectional analysis of observational data and does not support a causal claim. Trials requiring English-language proficiency differ systematically from those that do not, and unmeasured confounding remains possible. The positive and negative control analyses were intended to probe this possibility and behaved as predicted, but they cannot exclude residual confounding.

Targeted recruitment may confound these associations. Sponsors who add Spanish accommodation are plausibly recruiting where Hispanic/Latinx populations are concentrated, so documented language accommodation may be conflated with site selection. We examined this possibility by adjusting the enrollment models for US Census region and for the population share of the outcome-matched group across states containing trial sites. Trials requiring English-language proficiency were located in states with an average Hispanic/Latinx population share of 17.5%, compared with 19.5% for trials without the requirement. These measured geographic differences did not fully account for the associations: geographic adjustment left the requirement associations materially unchanged and attenuated the accommodation association, which remained substantial (**Supplemental Table 4**). Residual targeting at a finer geographic scale than the state cannot be excluded, because the registry records site states rather than catchment populations. The inverse association between documented language accommodation and the enrolled proportion of Asian and White participants may reflect the compositional structure of the race data and targeted recruitment. These associations therefore warrant cautious interpretation.

Finally, trials with posted results are not representative of all registered trials, so these estimates apply to trials with posted results[43]. Registry categories are also coarse: “Asian” spans subgroups that differ in language, culture, socioeconomic profile, and disease risk, which a single category may obscure[44,45].

## Conclusions

The prevalence of English-language proficiency requirements in clinical trials more than tripled between 2010 and 2023. These requirements were associated with lower enrolled proportions of Asian and Hispanic/Latinx participants. The less common practice of documenting language accommodation was associated with a higher enrolled proportion of Hispanic/Latinx participants. Digital and decentralized trials, promoted as vehicles of inclusion, had English-language requirements more often than conventional trials. Because these criteria are written by investigators, publicly recorded, and in principle amenable to revision, they may offer a tractable target for improving enrollment equity.

## AUTHOR CONTRIBUTIONS

DSK had full access to all the data in the study and takes responsibility for the integrity of the data and the accuracy of the data analysis. Concept and design: DSK, EY, FR, CTL, NS, AS. Acquisition, analysis, or interpretation of the data: AMT, LD, NS, PS, JWO, AJ, DSK. Registry adjudication: DSK and JWO. Drafting of manuscript: AMT, LD, DSK. Critical review of the manuscript: all authors. Statistical analysis: AMT, JWO, DSK. Supervision: DSK, EY, FR.

## DATA AND CODE AVAILABILITY

All primary data analyzed in this study are publicly available from ClinicalTrials.gov (https://clinicaltrials.gov). No individual participant data were used. All demographic variables were extracted from the aggregate baseline characteristic tables of the registry’s posted-results module. The analytic dataset, the complete set of classifier regular-expression patterns, the two-reviewer adjudication file used for classifier validation, and all analysis code are available at https://doi.org/10.5281/zenodo.22654518. The registry snapshot underlying the analysis was harvested in October 2026; because ClinicalTrials.gov is continuously updated, re-harvesting at a later date will not reproduce the cohort exactly, and the deposited analytic dataset is therefore the reproducible record.

## Supplemental Appendix 1. Verbatim regular expression classifier patterns

### English requirement

english[\s-]*speak(ing|er)?|able to (speak|read|understand|communicate)(in|\s)?\s*english|(speak|read|understand|communicate)s?\s+english|fluen(t|cy)\s+in\s+english|english\s+(proficiency|literacy)|proficient\s+in\s+english|native\s+english|english\s+as\s+a?\s*(first|primary)\s+language|(speak|read|write|understand|communicate)(,?\s*(speak|read|write|under stand|communicate))*\s*(,?\s*(and|or)\s*(speak|read|write|understand|communicate))?\s+(in\s+)?english|non[\s-]?english[\s-]?speak|not\s+(speak|read|understand)\s+english|unable\s+to\s+(speak|read|understand)\s+english|cannot\s+(speak|read|understand)\s+english

### Documented language accommodation

english\s+(or|/)\s+spanish|spanish\s+(or|/)\s+english|english\s+or\s+[a-z]+[\s-]?speaking Spanish mention (alternative exposure): spanish

Oral-medication capability (post hoc negative control)

able to swallow|swallow (tablets|capsules|pills|whole)|oral medication intake

### Digital modality

wearable|digital\s+health|accelerometer|mhealth|ehealth|activity\s+tracker|fitbit|actigraph|apple\ s*watch|garmin|smartwatch|continuous\s+glucose\s+monitor|\bcgm\b|pedometer|smartphone|m obile\s*app(lication)?|app-based|telehealth|telemedicine|decentrali[sz]ed|web-based|text\s*message|\bsms\b|virtual\s+(visit|trial|care)

### Covariate construction

Intervention class was derived from the registry interventions array: trials whose interventions included any BEHAVIORAL type were classified as “behavioral”; those including DRUG, BIOLOGICAL, or DEVICE were classified as “drug/biologic/device”; remaining trials were classified as “other”. Behavioral classification took precedence when both were present.

Therapeutic area was assigned by case-insensitive keyword search over the conditions array and the brief title, in fixed precedence order: oncology, mental_behavioral, cardiometabolic, infectious, neurological, respiratory, musculoskeletal, and other as the fallback when no pattern matched. Precedence is consequential because the keyword sets overlap: 2,302 trials (4.1%) match two or more area patterns, and stroke appears in both the cardiometabolic and neurological sets, where precedence assigns it to cardiometabolic. The first matching area was assigned, so a trial in more than one area received the first matching label.

Industry sponsorship was a binary indicator for a lead sponsor class of INDUSTRY; all other classes (NIH, other US federal, academic, other) formed the reference group.

Registered phase was a binary indicator for the presence of any phase designation in the registry phase field, because many behavioral trials have no registered phase.

Log enrollment and log sites were natural logarithms of one plus the actual (or, where actual was absent, estimated) enrollment count and the count of listed facilities, respectively.

Start year was the calendar year of the registry start date, mean-centered across the analytic cohort before entry into models. Digital or decentralized modality was flagged from the concatenation of the brief title and the brief summary text, matched against the term list above.

Outcome proportions were computed as the category count divided by the sum of all reported category counts for that measure, including Unknown or Not Reported. The known-race sensitivity denominator excluded Unknown or Not Reported from the denominator.

---

**Supplemental Table 1.** Confusion matrix for regular expression classifier validation against two physicians.

| **English language requirement** |  |  |  |
| --- | --- | --- | --- |
|  | **English requirement** | **No documented requirement** | **Total** |
| **RegEx Classifier +** | 15 (TP) | 1 (FP) | 16 |
| **RegEx Classifier -** | 2 (FN) | 132 (TN) | 134 |
| **Total** | 17 | 133 | 150 |
| **Documented language accommodation** |  |  |  |
|  | **Bilingual** | **No documented accommodation** | **Total** |
| **RegEx Classifier +** | 6 (TP) | 0 (FP) | 6 |
| **RegEx Classifier -** | 1 (FN) | 143 (TN) | 144 |
| **Total** | 7 | 143 | 150 |

Abbreviations: FN - false negative; FP - false positive; RegEx - regular expression; TN - true negative; TP - true positive;

**Supplemental Table 2.**
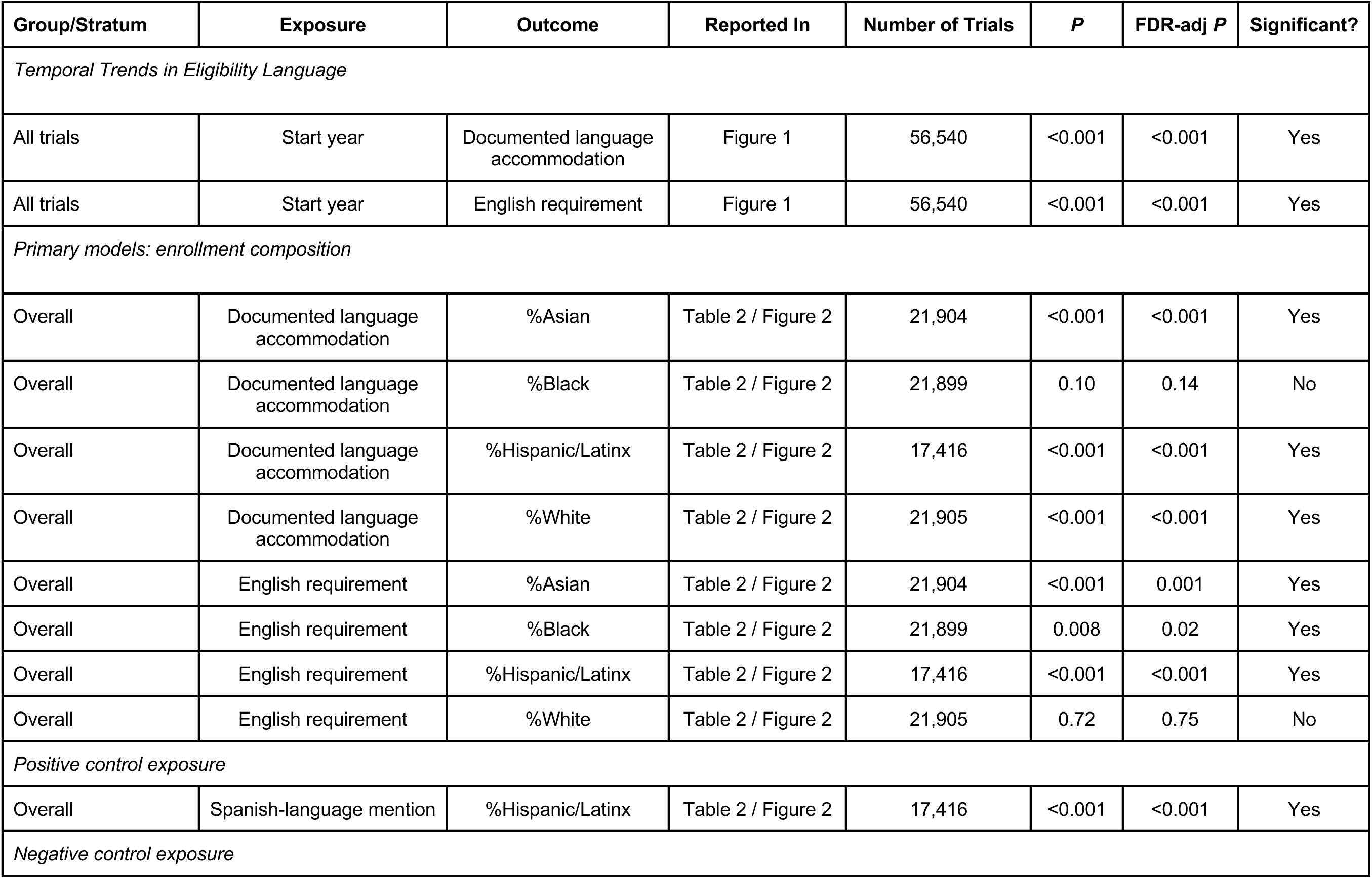
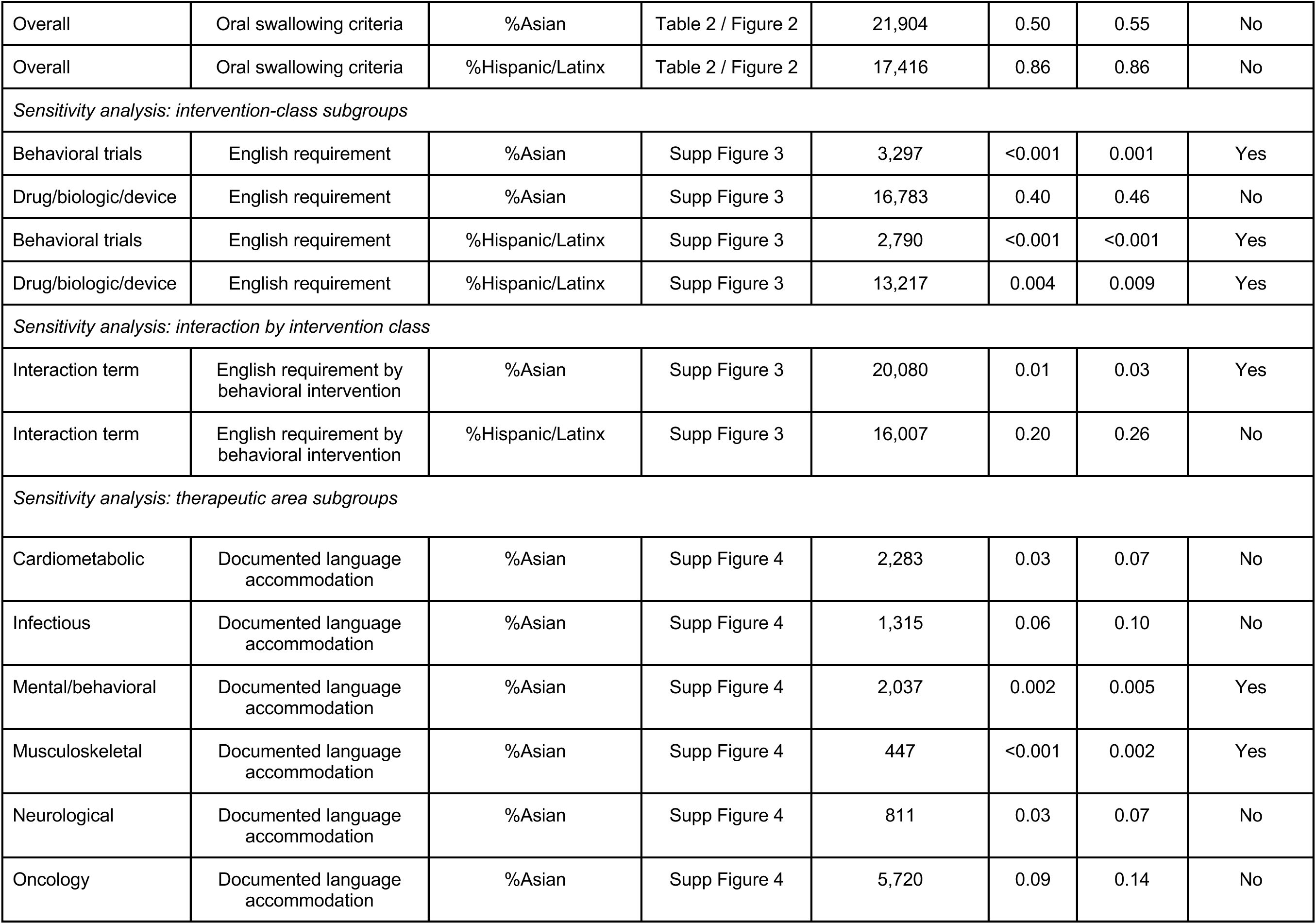
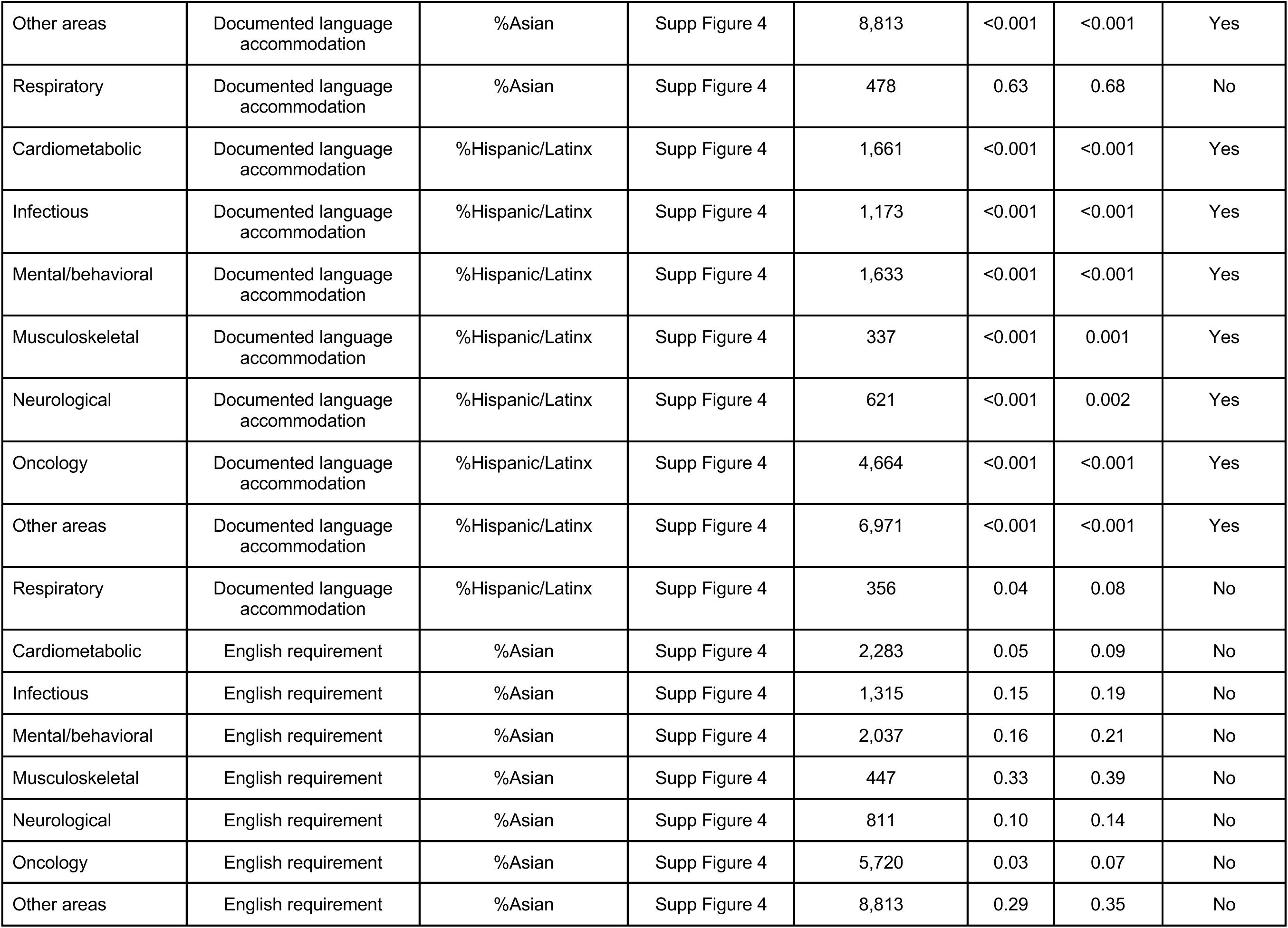
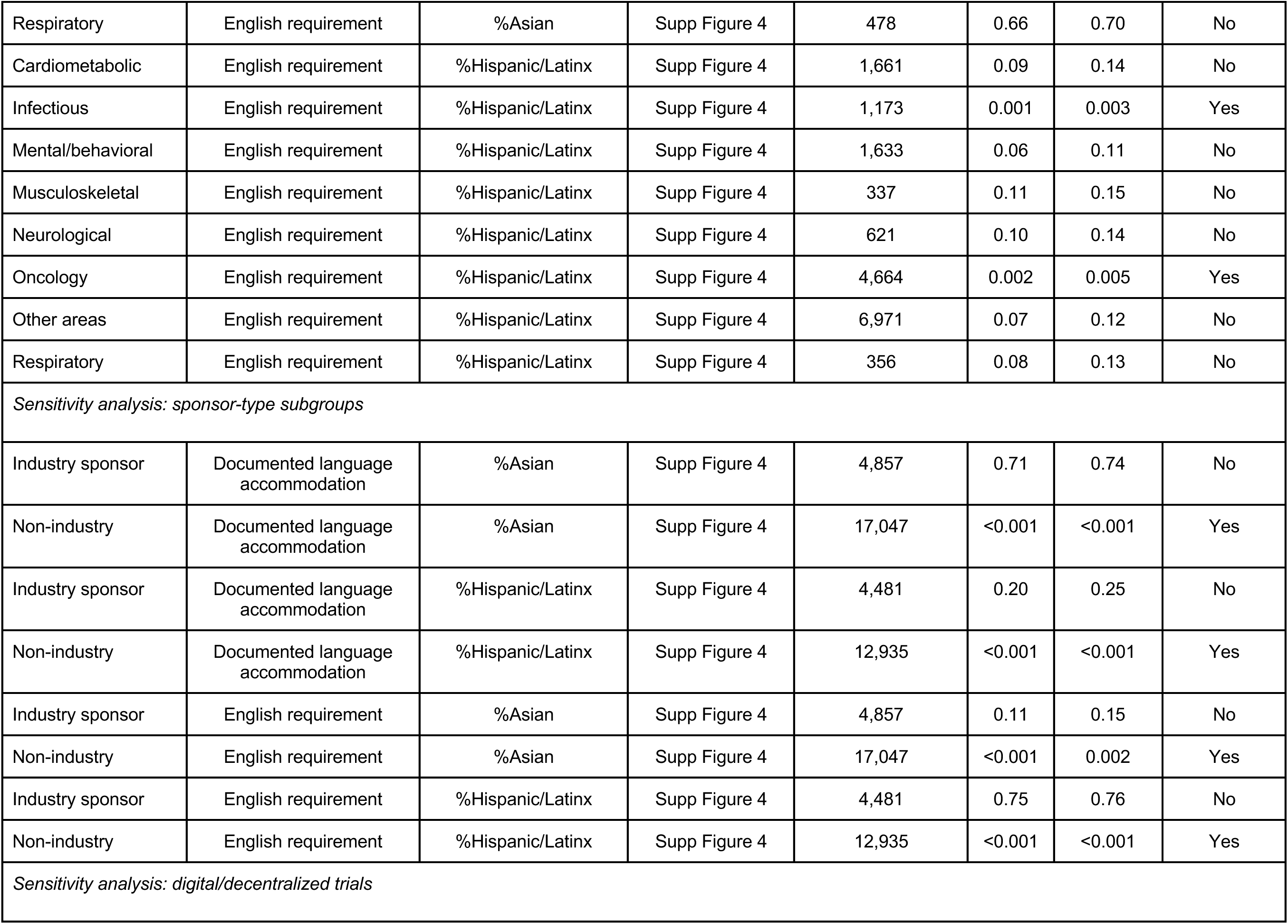
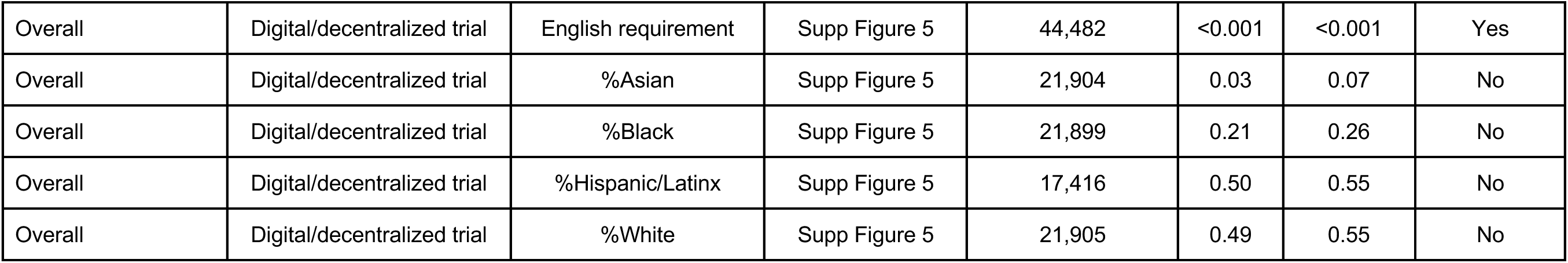
Complete inventory of every distinct statistical test reported in this article or in the eSupplement. Tests are grouped by type. A single Benjamini-Hochberg false discovery rate (FDR) correction is applied across all 64 tests, and every P value quoted in the text is taken from this table. Significance indicates an FDR-adjusted P value < 0.05.

**Supplemental Table 3.**
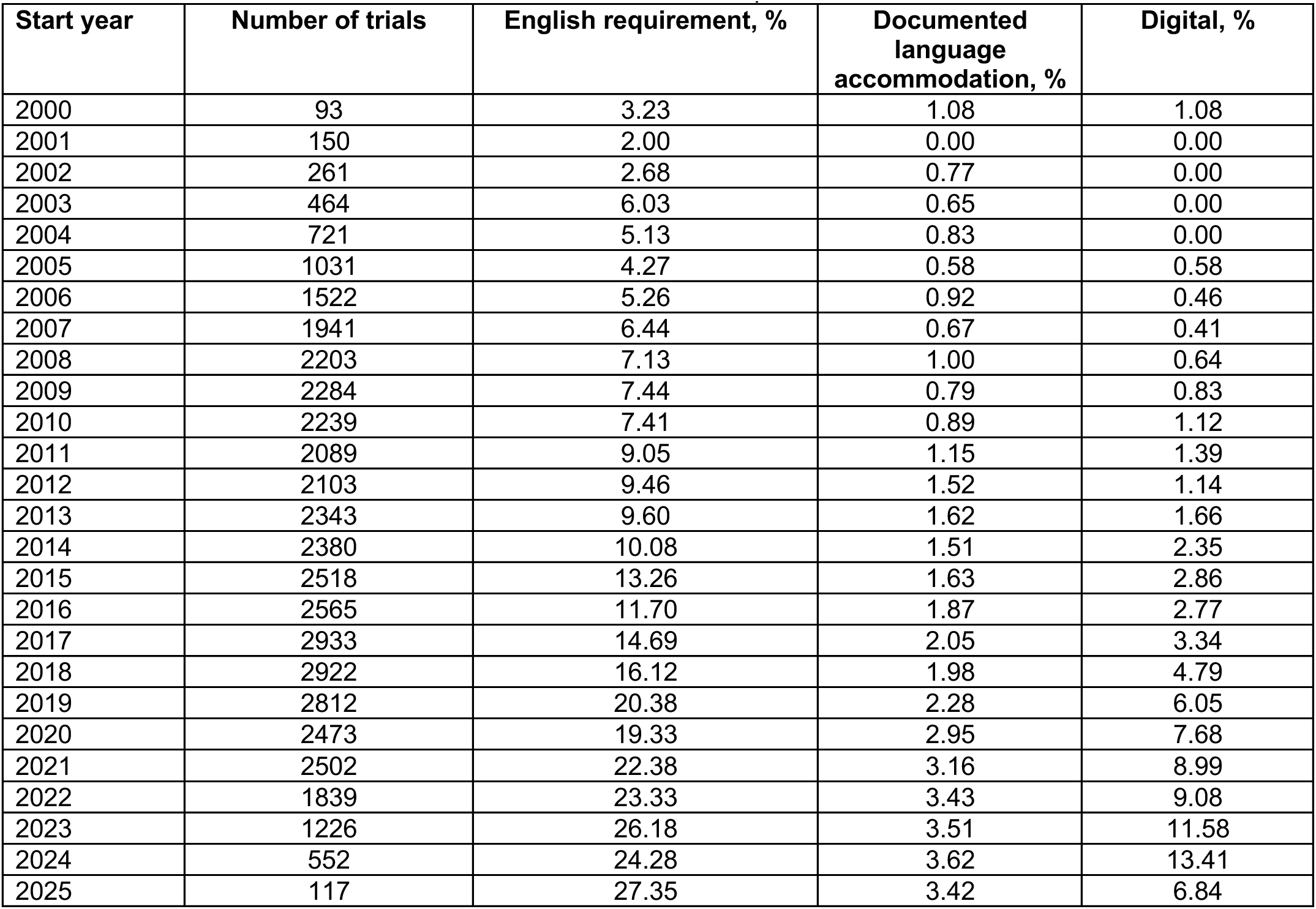
Year-by-year prevalence of each exposure and of the digital/decentralized trial modality (data underlying. **Figure 1).** Trial counts for 2024 (N=552) and 2025 (N=117) are far smaller than for 2016-2023 (N=1,226-2,933) because the cohort requires posted results; trials starting in these years are disproportionately still ongoing or within the results-submission window. Prevalence estimates for 2024-2025 should be interpreted with caution and were not used in the trend model.

| Start year | Number of trials | English requirement, % | Documented language accommodation, % | Digital, % |
| --- | --- | --- | --- | --- |
| 2000 | 93 | 3.23 | 1.08 | 1.08 |
| 2001 | 150 | 2.00 | 0.00 | 0.00 |
| 2002 | 261 | 2.68 | 0.77 | 0.00 |
| 2003 | 464 | 6.03 | 0.65 | 0.00 |
| 2004 | 721 | 5.13 | 0.83 | 0.00 |
| 2005 | 1031 | 4.27 | 0.58 | 0.58 |
| 2006 | 1522 | 5.26 | 0.92 | 0.46 |
| 2007 | 1941 | 6.44 | 0.67 | 0.41 |
| 2008 | 2203 | 7.13 | 1.00 | 0.64 |
| 2009 | 2284 | 7.44 | 0.79 | 0.83 |
| 2010 | 2239 | 7.41 | 0.89 | 1.12 |
| 2011 | 2089 | 9.05 | 1.15 | 1.39 |
| 2012 | 2103 | 9.46 | 1.52 | 1.14 |
| 2013 | 2343 | 9.60 | 1.62 | 1.66 |
| 2014 | 2380 | 10.08 | 1.51 | 2.35 |
| 2015 | 2518 | 13.26 | 1.63 | 2.86 |
| 2016 | 2565 | 11.70 | 1.87 | 2.77 |
| 2017 | 2933 | 14.69 | 2.05 | 3.34 |
| 2018 | 2922 | 16.12 | 1.98 | 4.79 |
| 2019 | 2812 | 20.38 | 2.28 | 6.05 |
| 2020 | 2473 | 19.33 | 2.95 | 7.68 |
| 2021 | 2502 | 22.38 | 3.16 | 8.99 |
| 2022 | 1839 | 23.33 | 3.43 | 9.08 |
| 2023 | 1226 | 26.18 | 3.51 | 11.58 |
| 2024 | 552 | 24.28 | 3.62 | 13.41 |
| 2025 | 117 | 27.35 | 3.42 | 6.84 |

**Supplemental Table 4.** Sensitivity of the primary associations to geographic adjustment. Each row reports the adjusted odds ratio (95% CI) for the same association under four specifications: no geographic adjustment; adjustment for US Census region of the trial’s sites; adjustment for the population share of the outcome-matched group across the trial’s site states, from the 2023 American Community Survey five-year estimates (table B03002) weighted by state population; and adjustment for both, which is the specification reported in Table 2. All models additionally include intervention class, therapeutic area, industry sponsorship, registered phase, total enrollment, number of sites, and trial start year.

| Exposure | Outcome | No geographic adjustment | + Census Region | + Site-state population share | + Both (reported) |
| --- | --- | --- | --- | --- | --- |
| English requirement | %Asian | 0.86 (0.78-0.94) | 0.85 (0.77-0.93) | 0.85 (0.78-0.93) | 0.85 (0.77-0.93) |
| English requirement | %Hispanic/Latinx | 0.85 (0.79-0.91) | 0.85 (0.79-0.91) | 0.86 (0.80-0.93) | 0.85 (0.79-0.91) |
| Language accommodation | %Hispanic/Latinx | 4.11 (3.59-4.70) | 3.78 (3.30-4.33) | 3.32 (2.90-3.81) | 3.23 (2.82-3.71) |
| Language accommodation | %Asian | 0.72 (0.61-0.86) | 0.62 (0.52-0.73) | 0.60 (0.51-0.71) | 0.59 (0.50-0.70) |

**Supplemental Figure 1.**
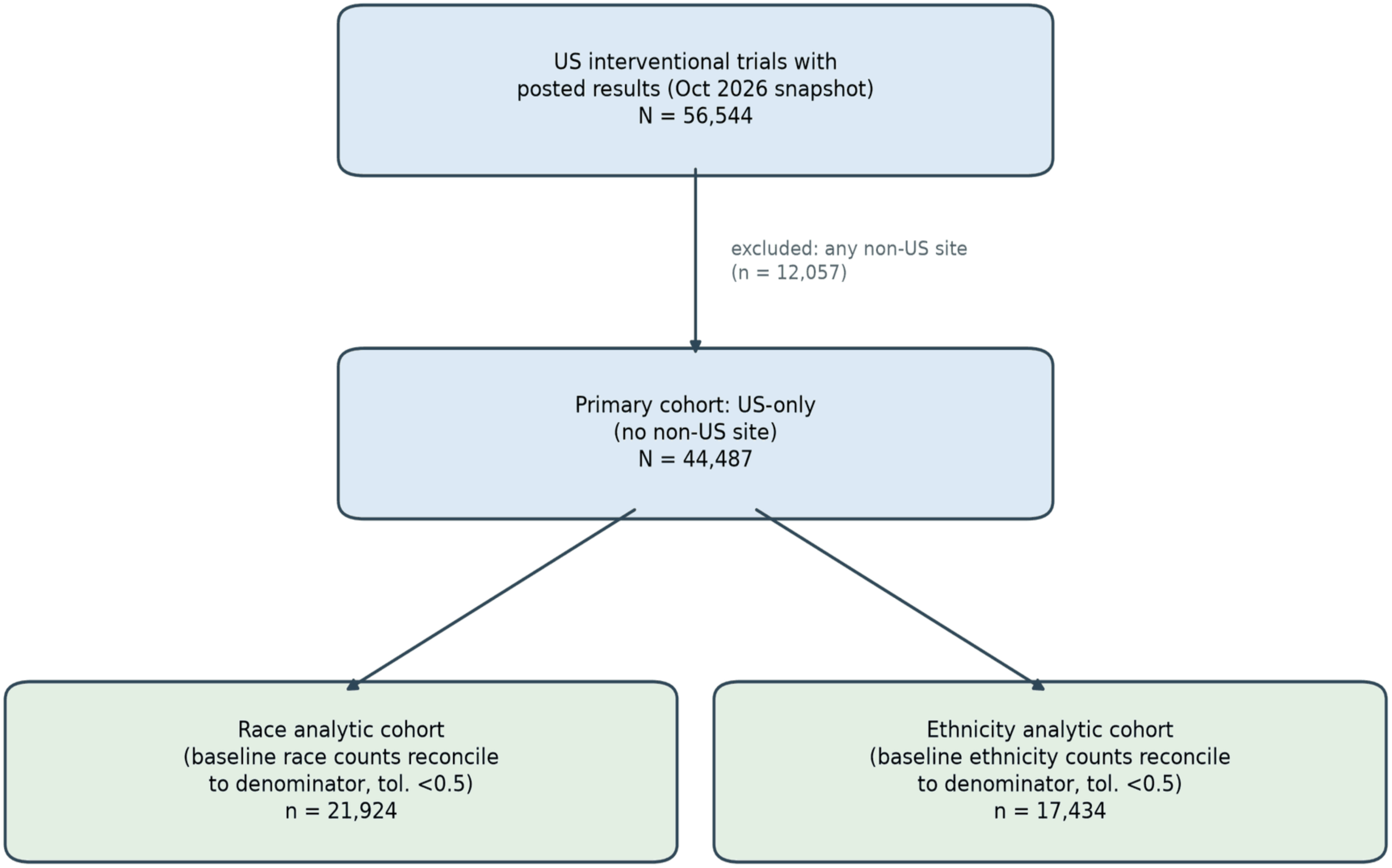
Cohort flow from the <u>ClinicalTrial.gov</u> registry records to the race and ethnicity analytic cohorts.

**Supplemental Figure 2.**
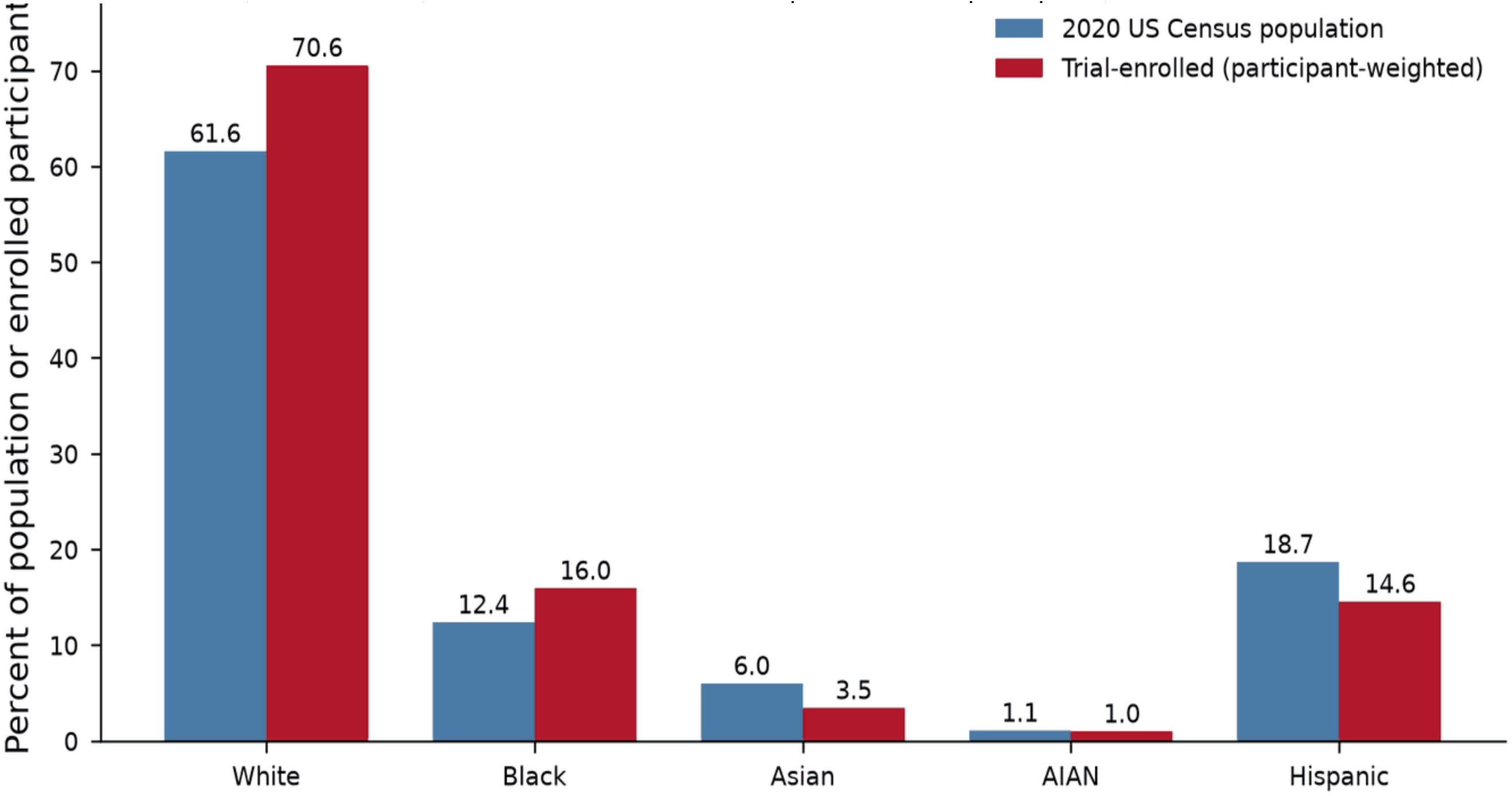
Participant-weighted enrolled composition (in US interventional trials with results) versus 2020 US Census population shares. AIAN=American Indian and/or Alaska Native. Census bars are 2020 race-alone shares with Hispanic or Latino origin reported separately, matching the two-question structure of the registry’s baseline module, so Hispanic or Latino participants are also counted in a race category on both sides. The registry denominators additionally contain participants whose race (6.6%) or ethnicity (9.2%) was not reported, for which the Census denominator has no counterpart; excluding them raises the ratios to 1.23 for White, 1.38 for Black, 0.62 for Asian and 0.86 for Hispanic or Latinx participants, without changing any direction.

**Supplemental Figure 3.**
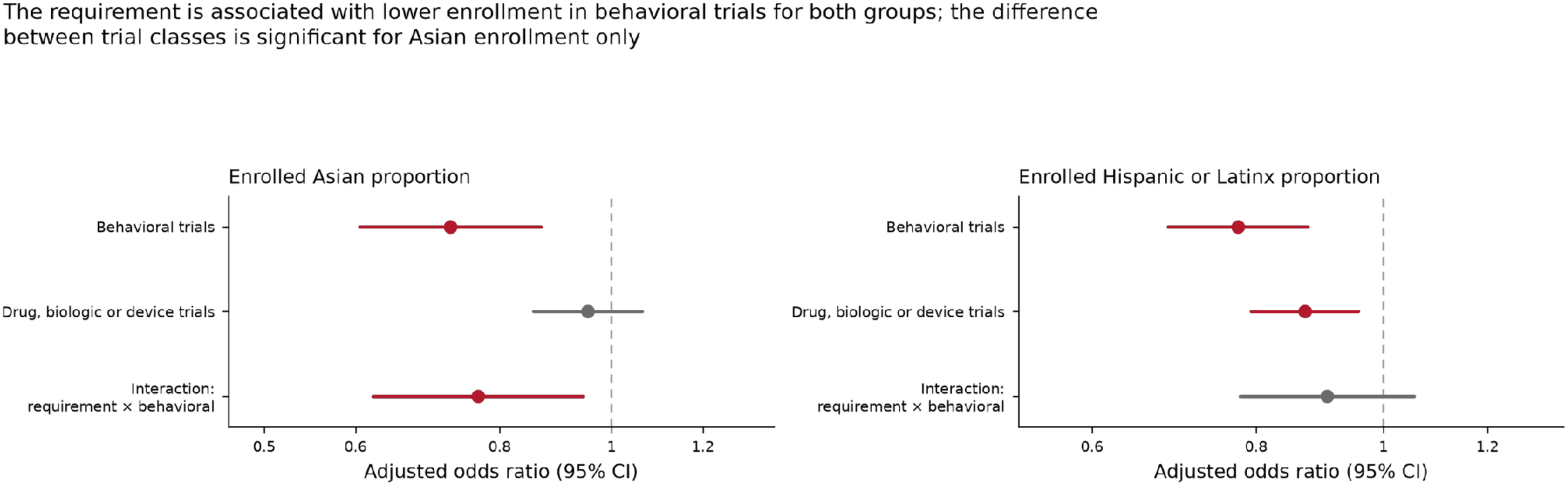
English-language proficiency requirement and enrolled proportion of Asian and Hispanic/Latinx participants, stratified by intervention class.

**Supplemental Figure 4.**
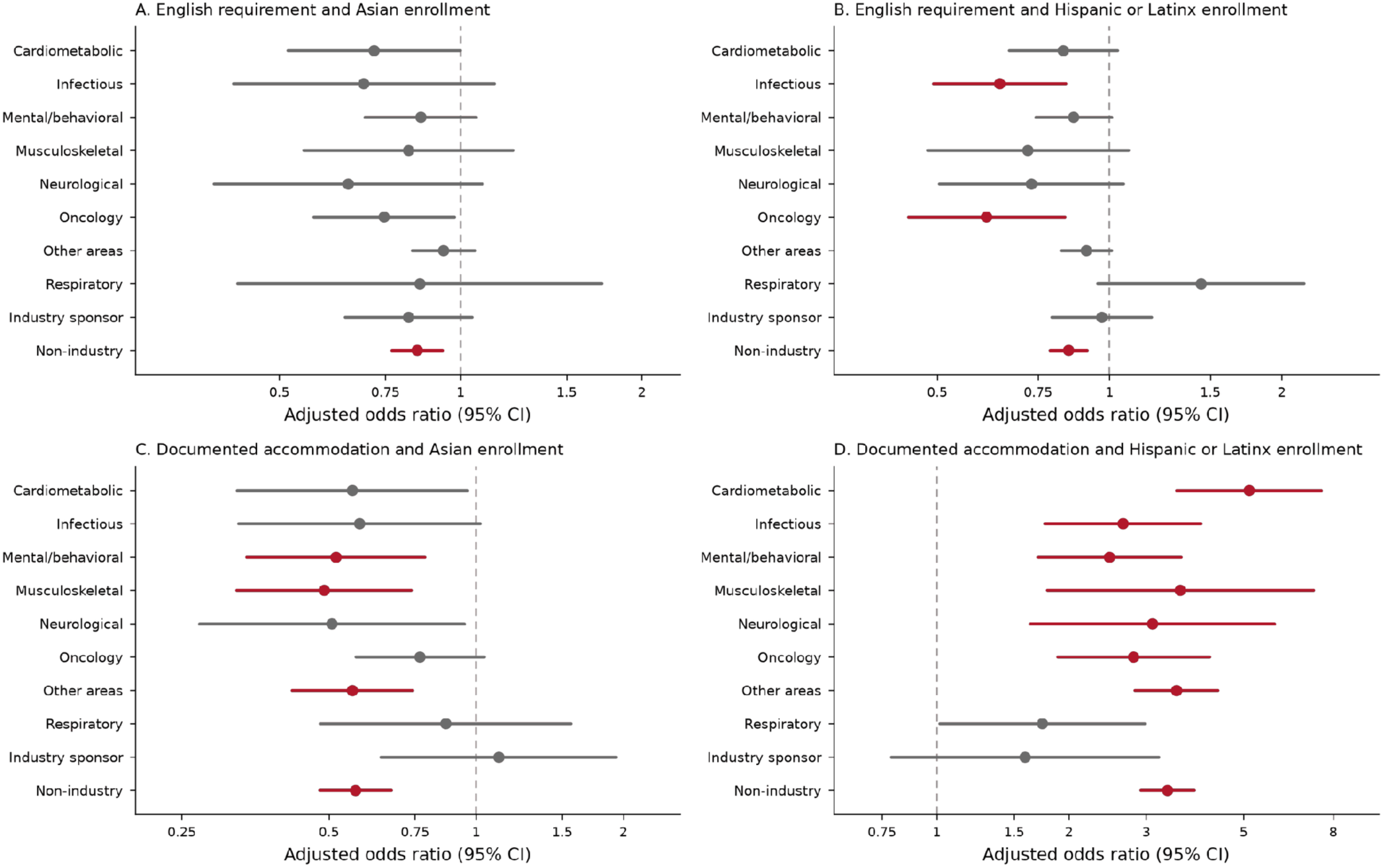
Association of English-language proficiency requirements and documented language accommodation with Asian and Hispanic/Latinx enrollment in clinical trials, stratified by therapeutic area and sponsor type. Each OR and 95% CI is the association between the exposure and enrolled composition among trials within the named stratum. The figure does not explicitly compare enrolled composition between strata. Red marks indicate an FDR-adjusted P<0.05 over the 73-test family, matching the significance column of eTable 2; the plotted confidence intervals are unadjusted 95% intervals, so an interval that excludes the null does not by itself establish significance after adjustment.

**Supplemental Figure 5.**
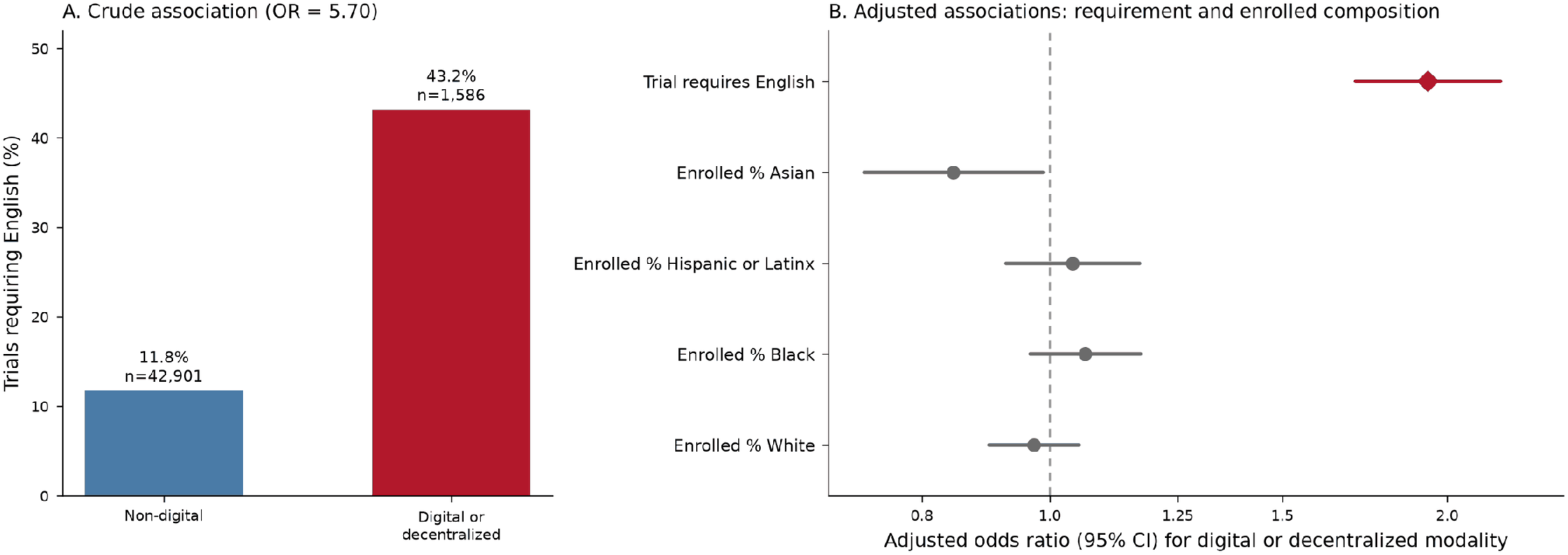
Digital or decentralized modality: crude association with English-language proficiency requirements (A) and adjusted associations with English-language proficiency requirements and enrolled racial and ethnic composition (B).

